# Knowledge of Sunscreen Usage and Skin Cancer Among Malaysian Medical Students – A cross sectional study

**DOI:** 10.1101/2023.01.28.23285149

**Authors:** Hazwani Nuruljannah binti Haris Fadzilah, Lee How Yea, Minduli Thirasaree Jayasree Dumingu Hewage, Fathima Salima Mohamed Azme, Htoo Htoo Kyaw Soe, Soe Moe, Mila Nu Nu Htay

## Abstract

**Background:** Excessive ultraviolet light (UV) can cause premature skin aging and potentially skin cancer. This study evaluated the knowledge, attitude of sunscreen, and skin cancer among Malaysian medical students.

**Methods:** This cross-sectional study was conducted from October 2022 to November 2022 among the clinical year medical students in a private medical university in Malaysia. The respondents were recruited by purposive sampling method. The content validated questionnaire was used to collect the data, and the data collection was done via online platform.

**Results:** There are a total of 117 responses that we collected through an online questionnaire via Google Forms. Among the respondents, 59.8% of the respondents reported of having a poor knowledge about sunscreen. However, 64.96% reported to have good knowledge about skin cancer. Approximately half of the respondents (48.7%) had a good attitude towards sunscreen. Females are more likely to use them compared to men (OR: 9.12, 95% CI: 3.52, 23.64) and there is a difference between ethnicity and the usage of sunscreen.

**Conclusion:** This study demonstrated limited knowledge of sunscreen among medical students. However, they are having better knowledge of skin cancer. Our results spotted the need for education about sunscreen among Malaysian medical students.

## Introduction

Initially, introduction of sunscreen to the population aims to prevent development of sunburns on skin, due to excessive exposure to sunlight. Sunburns are mainly caused by ultraviolet B (UVB) rays, however ultraviolet A (UVA) carries more damage to the skin (Hanrahan, 2012). It has been reported that unprotected exposure of the skin for five to ten minutes from the sun could be harmful to certain areas on the body which are more sensitive, namely the face, ears, and neck. UVA are more likely the culprit for long-term effects like production of age spots, photoaging and wrinkles. It acts by damaging the mitochondrial and nuclear DNA, causes mutation in genes that can lead to skin cancer, suppression of the immunity system, and photoallergic and phototoxic effects. UVB rays on the other hand is causes more direct effect such as sunburns, and pigmentations, which mainly prompts people to start using sunscreen (Korrapati et al., 2021). This being the reason why sunscreen was initially created to defend against mainly UVB, which correlates to direct exposure of the sunlight.

According to the reports of research and evidence, sunscreen products that protect against both UVA and UVB increasing nowadays (Hanrahan, 2012). A study carried out in a public university in Malaysia compared the knowledge and attitude of sunscreen usage between final year medical and pharmacy students. The findings concluded that pharmacy students had higher attitude towards the practice of sunscreen compared to medical students due to the fact that they had been exposed to lectures on nutraceuticals and cosmeceuticals, which were particular subjects on sunscreen (Awadh et al., 2016). A similar study was done by Qin Jian Low, comparing the usage of sunscreen among pharmacists and doctors in Malaysia, also concluded that pharmacists had a higher rate of sunscreen usage compared to doctors. Among the pharmacists, women had more awareness for sunscreen protection (Low et al., 2021). However, despite high awareness of using sunscreen as their daily routine, majority of the research participants were not aware of the UV rays that caused a higher risk for skin cancer (Low et al., 2021). Skin cancer is a category of illnesses distinguished by abnormal cell growth and mutation in the skin’s epidermal layer (Al-Atif, 2021). Over the past few decades, both non-melanoma and melanoma skin cancer rates have increased globally (WHO, 2017). Currently, 132,000 cases of melanoma and 2 to 3 million cases of non-melanoma skin cancer are reported annually worldwide (WHO, 2017). According to statistics from the Skin Cancer Foundation, one in every three malignancies diagnosed are skin cancers, and one in every five Americans will have skin cancer at some point in their lives (WHO, 2017). Skin cancer was the tenth most prevalent cancer in Malaysia, according to the third report from the National Cancer Registry (2003-2005) (Affandi, 2018). However, in contrast to other malignancies, skin cancer has not gotten much attention in Malaysia, and the general population has limited awareness on skin cancer. There are two major types of skin cancer: melanoma, non-melanoma. Melanoma type of tumor appears very rarely. It develops on the skin cell called melanocytes. Non-melanoma skin cancer is a common term used to describe Basal Cell Carcinoma (BCC) and Squamous Cell Carcinoma (SCC) (Liu-Smith et al., 2017; Qadir, 2016). Melanoma is a kind of skin cancer that, if not found and treated at an early stage, can be deadly (Diao & Lee, 2013). It has the ability to metastasize and become life-threatening (Diao & Lee, 2013). Melanoma also known as malignant melanoma or cutaneous melanoma (CancerResearchUK, 2020).^11^ Sun exposure is considered to be a significant environmental risk factor for melanoma, basal cell carcinoma (BCC) and squamous cell carcinoma (SCC). The formation of SCC is thought to be caused by cumulative solar damage (AIHW, 2022; Gallagher et al., 1995; Kricker et al., 1994; Zanetti et al., 2006), while the development of BCC and melanoma appears to be caused by a combination of cumulative and intermittent solar damage (AIHW, 2022; CancerResearchUK, 2020; Diao & Lee, 2013; Gallagher et al., 1995; Kricker et al., 1994; Lee et al., 2013; Zanetti et al., 2006).

Skin cancer may affect anybody, however those with light (fair) skin tones are more likely to get it. Although it can appear anywhere on the body, it is most frequently observed on areas of the face, arms, or hands that are constantly exposed to sunlight. The most typical warning signs and symptoms of skin cancer include changes in a mole’s size, colour, or form; seeping or bleeding from a mole; a mole that feels itchy; hard, lumpy, or swollen; and a growth or sore that won’t go away (NationalCancerInstitute, 2016). Study done in a Turkish University shows that the knowledge of skin cancer is shown better to those that studied regarding the topic in their respective departments, which also in general, improves their behaviour towards skin cancer (Uğrlu et al., 2016). Exposure to sun radiation is the main cause of skin cancer, along with the skin’s vulnerability to sunlight’s harmful effects, including a pale complexi on and a propensity to burn, blister, or freckle in the sun (Awadh et al., 2016; Benjamin & Ananthaswamy, 2007). The skin undergoes significant intrinsic and extrinsic changes as a result of exposure to the sun. Regular use of sunscreen can help prevent a number of photoaging symptoms, including DNA and cell damage that leads to sagging, wrinkles, and photocarcinogenesis (Sander et al., 2020).

Previous research was conducted about sunscreen usage, attitude and knowledge, or prevention of skin cancer in various countries. However, to the best of our knowledge, there is no study as of date, done in Malaysia among medical students, to assess their knowledge and attitude towards sunscreen usage and its association with their knowledge of skin cancer. This study aimed to assess knowledge, attitude, practice of sunscreen and knowledge of skin cancer among undergraduate medical students in a private medical university in Malaysia.

## Methods

### Study setting and population

This cross-sectional study was conducted from October to November 2022 among the clinical year medical students in a private medical university in Malaysia. The medical students studying in the clinical were recruited in this study. The sample size was calculated using epi info calculator with an expected frequency of 36.6% sunscreen utilization (Awadh et al., 2016), with 7% margin of error and 95% confidence level. While taking into consideration the non-response rate 10%, the final estimated sample size was 117 students.

The sampling method used in this study was non-probability purposive sampling. The inclusion criteria were medical students (MBBS, either male or female, both international and local students, who choose to voluntarily agree to participate in the study and voluntarily provided informed consent to participate in this study. The exclusion criteria were pre-clinical year students, students studying in other programme, and those who do not provide informed consent.

### Data Collection

Data for this research was collected via an online questionnaire that was distributed among the respondents through social media platforms such as WhatsApp and Instagram. The questionnaire was prepared based on the previous studies (Awadh et al., 2016; Glanz et al., 2022; Memon et al., 2019; Sander et al., 2020) and content validated by using the ratings of six experts. The questionnaire included five sections which consisted of the demographics, practices of sunscreen utilization, knowledge about sunscreen, attitudes toward sunscreen and knowledge about skin cancer.

The demographics section (Section I) included age, gender, nationality, ethnicity, academic year, accommodation status, whether you are aware of the skin type, and to inquire if there is any family or personal history of skin cancer. Section II of the questionnaire, which focuses on the practices of sunscreen utilization was assessed with nine multiple-choice questions. Section III, practices regarding sunscreen application among medical students were inquired in the form of fifteen questions and the responses were recorded as “yes/no/do not know”. Section IV of the questionnaire focused on the attitude towards sunscreen among medical students and included eight items recorded with five-point Likert Scale. The final section (V) of the questionnaire was to assess the medical students’ knowledge regarding skin cancer. This section included ten questions in the form of close-ended questions.

### Data Analysis

Data was analyzed by using Epi Info (version 7.2). Descriptive analysis was conducted for the demographic variables. Inferential statistical analysis, Chi-square test, was used for hypothesis testing in the study and draw conclusions regarding the proper knowledge about sunscreen and skin cancer amongst medical students.

### Ethical Consideration

Prior to the study’s onset, all respondents received a full overview of the goal and action plan of the study. The respondents were then provided with an informed consent, where they were given the opportunity to decide on participating in the study. Moreover, the respondents were given the assurance that anonymity would be maintained, and data would be used for research purposes only. The participation was entirely voluntary. Even after signing the informed consent form, they were fully permitted to withdraw the study at any time. The ethics approval to conduct this study was granted by the Research Ethics Committee, Faculty of Medicine, Manipal University College Malaysia (MUCM), Malaysia.

## Results

A total of 117 students participated in this study. Table 1 shows the sociodemographic characteristics of the respondents. Among these responses, the majority (70.1%) were females and 68.4 % were below 22 years of age. Malay population represented 15.4 %, the Chinese by 26.5 %, Indian population by 31.6% and the other ethnicities by 26.5%. The majority (84.6%) of respondents were aware of their skin type and 1.7% of respondents reported of having a family or personal history of skin cancer (Table 1).

**Table 1.**
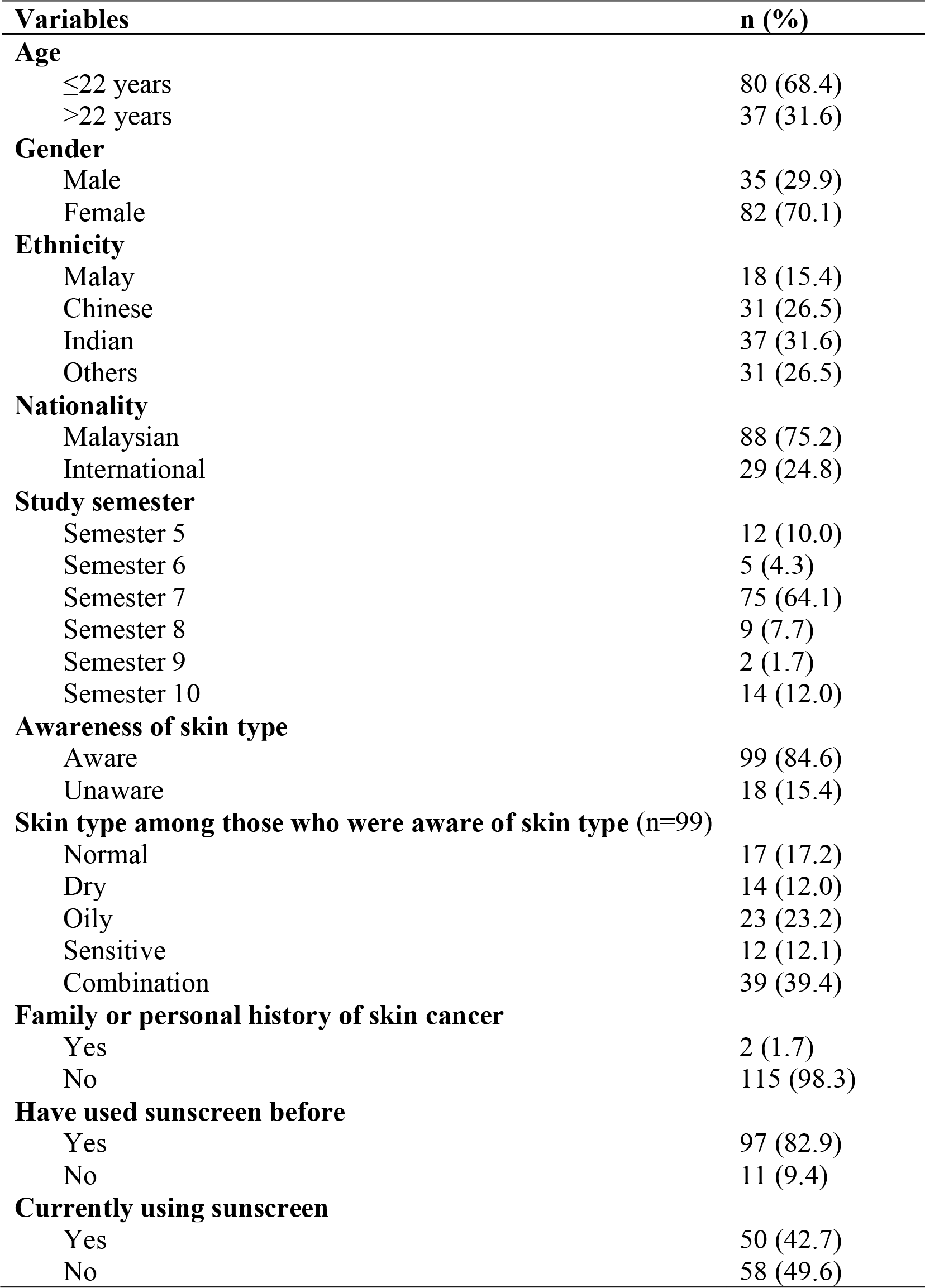
Sociodemographic characteristic of respondents (n=117)

| **Variables** | **n (%)** |
| --- | --- |
| **Age** |  |
| ≤22 years | 80 (68.4) |
| >22 years | 37 (31.6) |
| **Gender** |  |
| Male | 35 (29.9) |
| Female | 82 (70.1) |
| **Ethnicity** |  |
| Malay | 18 (15.4) |
| Chinese | 31 (26.5) |
| Indian | 37 (31.6) |
| Others | 31 (26.5) |
| **Nationality** |  |
| Malaysian | 88 (75.2) |
| International | 29 (24.8) |
| **Study semester** |  |
| Semester 5 | 12 (10.0) |
| Semester 6 | 5 (4.3) |
| Semester 7 | 75 (64.1) |
| Semester 8 | 9 (7.7) |
| Semester 9 | 2 (1.7) |
| Semester 10 | 14 (12.0) |
| **Awareness of skin type** |  |
| Aware | 99 (84.6) |
| Unaware | 18 (15.4) |
| **Skin type among those who were aware of skin type (n=99)** |  |
| Normal | 17 (17.2) |
| Dry | 14 (12.0) |
| Oily | 23 (23.2) |
| Sensitive | 12 (12.1) |
| Combination | 39 (39.4) |
| **Family or personal history of skin cancer** |  |
| Yes | 2 (1.7) |
| No | 115 (98.3) |
| **Have used sunscreen before** |  |
| Yes | 97 (82.9) |
| No | 11 (9.4) |
| **Currently using sunscreen** |  |
| Yes | 50 (42.7) |
| No | 58 (49.6) |

Among the respondents, 47 (40.2%) had good knowledge and 70 (59.8%) had poor knowledge about sunscreen. 82.05% of the respondents have gained knowledge about skin cancers from lecture class, 67.52% by internet, 41.03% by social media, 32.48% by books, 29.91% by health professionals, 17.95% by relatives or friends. The percentage of the respondents who answered individual correct items are reported in Table 2.

**Table 2.** Knowledge about sunscreen among the respondents (n=117)

| **Item** | **Frequency of correct answers (%)** |
| --- | --- |
| Sunscreen prevents sunburn | 96 (82.1) |
| Sunscreen prevents skin cancer | 83 (70.9) |
| Sunscreen prevents skin ageing | 67 (57.3) |
| Sunscreen reverses signs of ageing | 23 (19.7) |
| Sunscreen needs a half hour to be absorbed by skin to work best | 69 (59.0) |
| Sun Protective Factor (SPF) of 15 or higher allows people to stay outside for 3 hours without worrying about getting a burn | 29 (24.8) |
| Best type of sunscreen is protection against only UVA rays | 40 (34.2) |
| Sunscreen should be reapplied every 2 hours | 63 (53.8) |
| Sunscreen should be applied when attending lectures, working in hospital or any indoor activities | 55 (47.0) |
| Sunscreen should be applied when attending occasions at night | 75 (64.1) |
| Sunscreen application can cause vitamin D deficiency | 60 (51.3) |
| Minimum of SPF 15 & broad-spectrum sunscreen to reduce risk of skin cancer | 64 (54.7) |

The responses on attitudes towards sunscreen are reported in Table 3. Approximately half (48.7%) of the respondents revealed positive attitudes towards sunscreen, while 51.3% reported of poor attitudes.

**Table 3.** Attitudes towards sunscreen among medical students (n=117)

| **Item** | **Strongly Disagree** | **Somewhat Disagree** | **Neutral** | **Somewhat Agree** | **Strongly Agree** |
| --- | --- | --- | --- | --- | --- |
|  | **n (%)** | **n (%)** | **n (%)** | **n (%)** | **n (%)** |
| Sunscreen ingredients are toxic | 24 (20.5%) | 40 (34.8%) | 46 (39.3%) | 5 (4.3%) | 2 (1.7%) |
| Most sunscreen is full of harmful chemical | 26 (22.2%) | 38 (32.5%) | 38 (32.5%) | 13 (11.1%) | 2 (1.7%) |
| Sunscreen isn't safe to use | 42 (35.9%) | 41 (35.0%) | 33 (28.2%) | 0 (0%) | 1 (0.9%) |
| Sunscreen lotions are too expensive | 9 (7.7%) | 18 (15.4%) | 40 (34.2%) | 43 (36.8%) | 7 (6.0%) |
| Remembering to carry sunscreen when you go out is inconvenient | 18 (15.4%) | 20 (17.1%) | 26 (22.2%) | 39 (33.3%) | 14 (12.0%) |
| Sunscreen has an unpleasant smell | 20 (17.1%) | 32 (27.4%) | 36 (30.8%) | 23 (19.7%) | 5 (4.3%) |
| It is a hassle to reapply sunscreen | 12 (10.3%) | 13 (11.1%) | 29 (24.8%) | 46 (39.3%) | 16 (13.7%) |
| You're concerned about the safety of ingredients in sunscreen | 16 (13.7%) | 20 (17.1%) | 40 (35.1%) | 31 (26.5%) | 9 (7.7%) |

A total of 64 respondents (54.7%) reported that they always or frequently used sunscreen every time they went out in daylight. Approximately half (47.9%) of the respondents applied sunscreen 30 minutes before going out. The details of the sunscreen utilization practice are reported in Appendix Table 1. The association between the demographic characteristics of the respondents and sunscreen utilization are reported in Table 4. Female were more likely to utilize the sunscreen compared to male respondents (OR: 9.12, 95% CI: 3.52, 23.64). There was no significant association between age, ethnicity, and sunscreen utilization among the respondents (Table 4).

**Table 4.** association between the demographic characteristics of the respondents and sunscreen utilization (n=117)

| Variables | Utilization of sunscreen |  |  | OR | 95% CI | P value |
| --- | --- | --- | --- | --- | --- | --- |
|  | Always or frequent utilization<br>n (%) | Rarely or never utilization<br>n (%) |  |  |  |  |
| Age (years) |  |  |  |  |  |  |
| >22 | 20 | 17 | 37 | Reference |  |  |
| ≤ 22 | 44 | 36 | 80 | 1.039 | 0.475, 2.271 | 0.924 |
| Gender |  |  |  |  |  |  |
| Male | 7 | 28 | 35 | Reference |  |  |
| Female | 57 | 25 | 82 | 9.12 | 3.52, 23.64 | <0.001 |
| Ethnicity |  |  |  |  |  |  |
| Malay | 12 | 6 | 18 | Reference |  |  |
| Chinese | 14 | 17 | 31 | 0.41 | 0.12, 1.38 | 0.146 |
| Indian | 23 | 14 | 37 | 0.82 | 0.25, 2.68 | 0.745 |
| Others | 15 | 16 | 31 | 0.47 | 0.14, 1.57 | 0.215 |

Table 5 represents data on knowledge of skin cancer among students who frequently utilize sunscreen and those who rarely or never utilize sunscreen. The finding reveals that there was no statistically significant association between the skin cancer knowledge and utilization of sunscreen (Table 5).

**Table 5.**
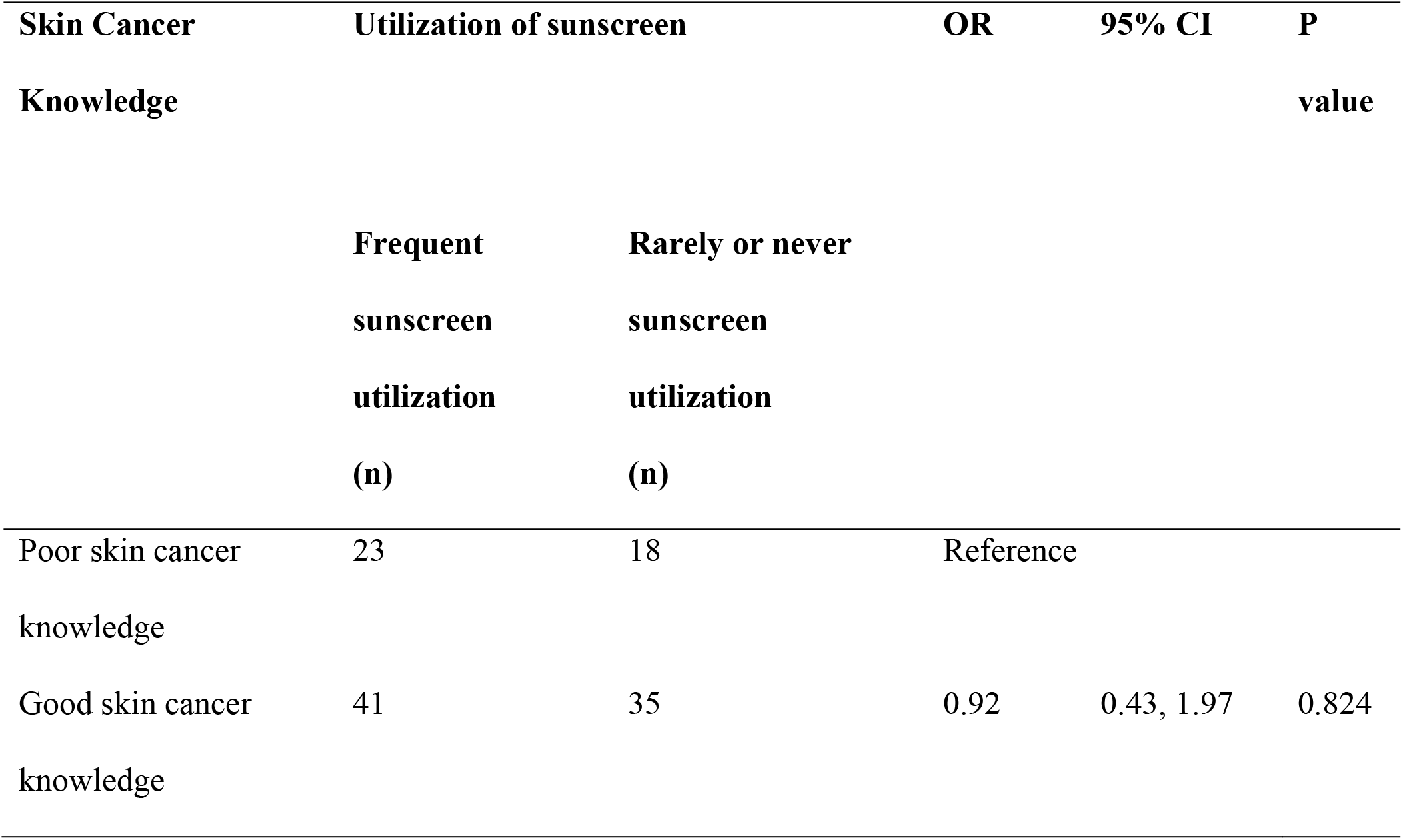
Respondents’ sunscreen utilization practice and their knowledge of skin cancer (n=117)

## Discussion

This cross-sectional study was carried out among undergraduate medical students to investigate the knowledge of sunscreen usage and skin cancer, to investigate the association of age, gender, and ethnicity towards sunscreen usage, and association of skin cancer knowledge and utilization of sunscreen. In this study we found out that among 117 respondents of clinical year medical students, only 54.7% of the students have a good attitude towards sunscreen usage. This is almost similar to a study done on medical students in Karachi, where only 60% are reported to have a good attitude towards sunscreen usage (Memon et al., 2019). Another cross-sectional study conducted in Indonesia reported that only 50% of the respondents have a good practice of applying sunscreen for their daily routine (Almuqati et al., 2019). It could be correlated with the knowledge of sunscreen usage in this study, only 40.2% of our students scored good knowledge. Medical students tend to have lower knowledge regarding sunscreen compared to other departments such as pharmacy students, as mentioned before, where medical students did not specifically study regarding sunscreens, nor did they have subjects such as nutraceuticals and cosmeceuticals (Awadh et al., 2016).

Female respondents in this study were more likely to utilize sunscreen compared to male respondents. Women also tend to have more awareness and concern regarding the danger of sun exposure, or due to the fact that they are more concerned about their image, which leads to higher knowledge and attitude towards sunscreen (Memon et al., 2019). This might be also related to the Asian belief where they considered being fair defines beauty, that makes them more compliant to use sunscreen (Low et al., 2021). This is proven in our study where the majority of the reason that prompts them to apply sunscreen is to avoid tanning. Aside from usage of sunscreen, it is also recommended by experts to practice other measures such as using protective attires, usage of widely brimmed hats, and staying in the shade to protect self from sun exposure (Hall et al., 1997).

Moreover, aside from protection against the sun exposure, production of naevi on the sun exposed skin that can predispose to melanoma can also be reduced with the application of sunscreens (Pincus et al., 1991). Our respondents, have generally good idea on when is the appropriate timing to apply sunscreen, as 47.9% of the respondents applies sunscreen 30 minutes before going out. This is as recommended by the Skin Cancer Org. where it is best to apply the sunscreen 30 minutes before going out to allow the sunscreen to set into the skin. It also should be reapplied after every 2 hours after exposure to the sun, or immediately after activities such as swimming or excessive sweating (SkinCancerFoundation, 2022). However, the majority of our respondents did not practice sunscreen reapplication. Respondents’ knowledge score of skin cancer was mostly good, and 82% of the respondents got their knowledge regarding skin cancer from lecture classes held by the university. If this is compared to studies from the Western countries, where majority of the respondents have awareness that skin cancer can be caused by sun exposure, but only a small percentage of 29% can identify the behavior that reduce the risk of developing skin cancer correctly (Almuqati et al., 2019). However, many of the respondents also agreed that the internet and social media had fueled up their knowledge regarding this topic. This was also agreed by many as they saw that social media platform could play a huge role in awareness of information regarding healthcare for the public. In regard to skin cancer, dermatologists not only chose this platform to give and spread awareness of the importance of using sunscreen, but they also use it as a mean to promote professional opinions and promote more patient-doctor interactions (De La Garza et al., 2021). Our cross-sectional study shows that a majority of the respondents (59.8%) had poor knowledge with regards to sunscreen. However, 65.0% of the respondents had good knowledge about skin cancer. Approximately half of the respondents (48.7%) had a good attitude towards sunscreen. Females are more likely to use them compared to men and there is a difference between ethnicity and the usage of sunscreen. The knowledge on sunscreen should be improved among medical students in Malaysia via education talk or lectures especially to the male student population. Furthermore, this current study might expand in various ways and will be of effective use in the future to those who would want to conduct in-depth investigations regarding the knowledge of skin cancer and sunscreen usage.

## Data Availability

All data produced in the present study are available upon reasonable request to the authors.

## Acknowledgements

First of all, we would like to give thanks to all of the respondents who volunteered in our study. We also wish to extend our heartfelt gratitude to the Vice Chancellor Dean Professor Dr. Adinegara Lutfi Abas, Dean Professor Dr. Jayakumar Gurusamy, and higher management from Manipal University College Malaysia. Apart from that, we would also like to thank the Research Ethics Committee, Faculty of Medicine, Manipal University College Malaysia, for approving our research.

## Authors contribution

We confirm that each author substantially contributed to the intellectual content of the paper and accepts public responsibility for that content.

## Funding

This research received no specific grant from any funding agency in the public, commercial, or not-for-profit sectors.

## Conflicts of interest

The authors declare that they have no competing interests.

## Appendix

**Appendix Table 1.**
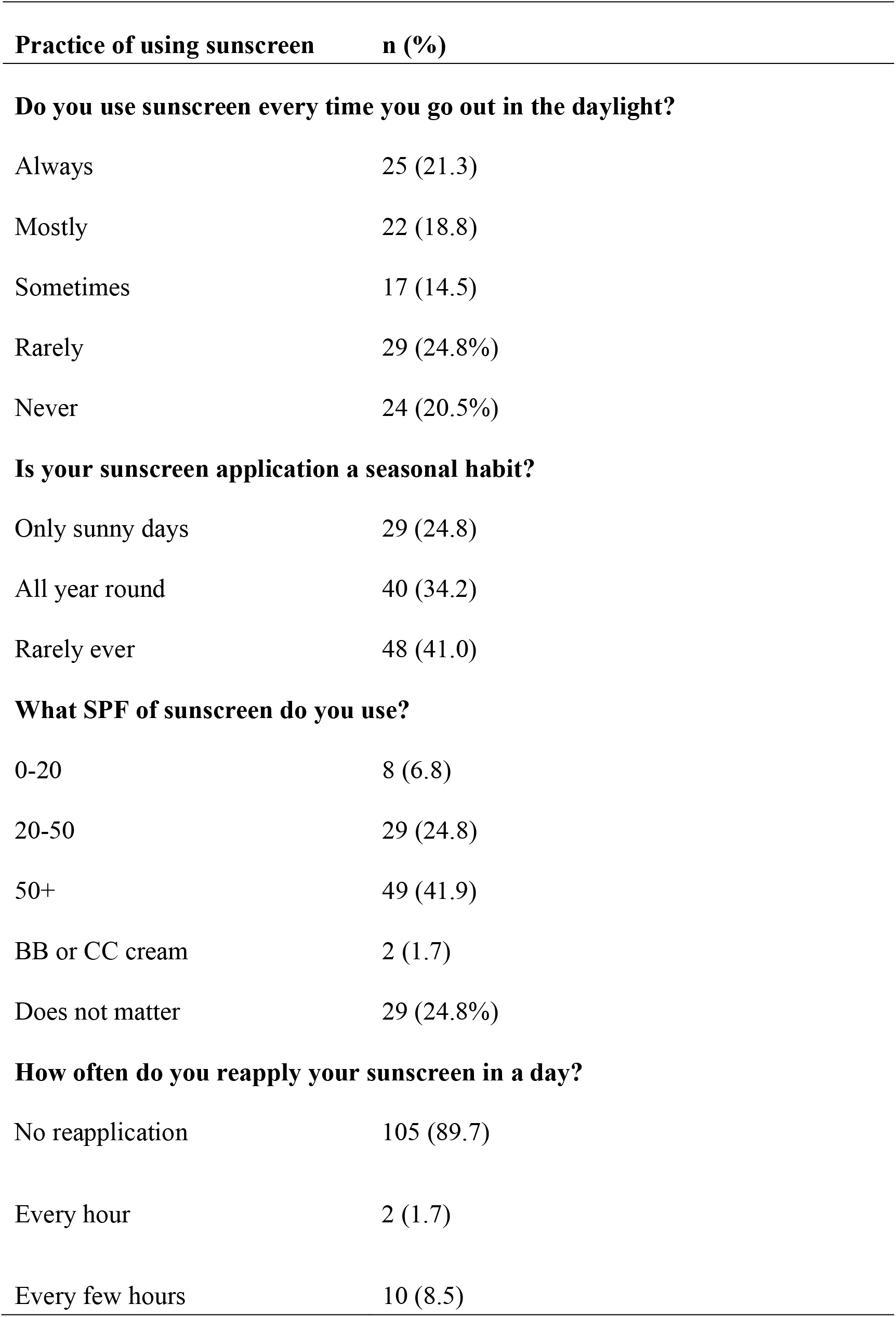
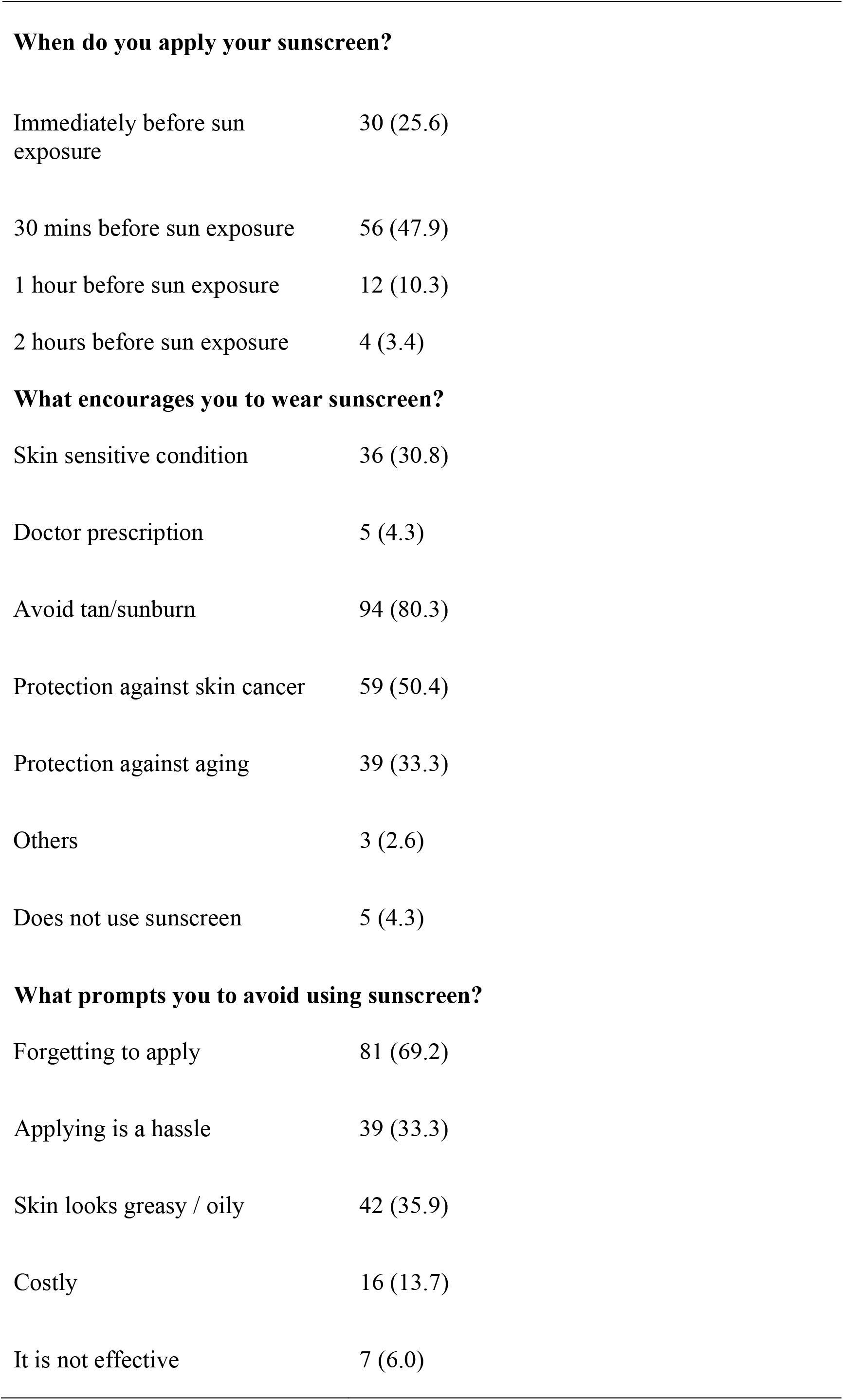
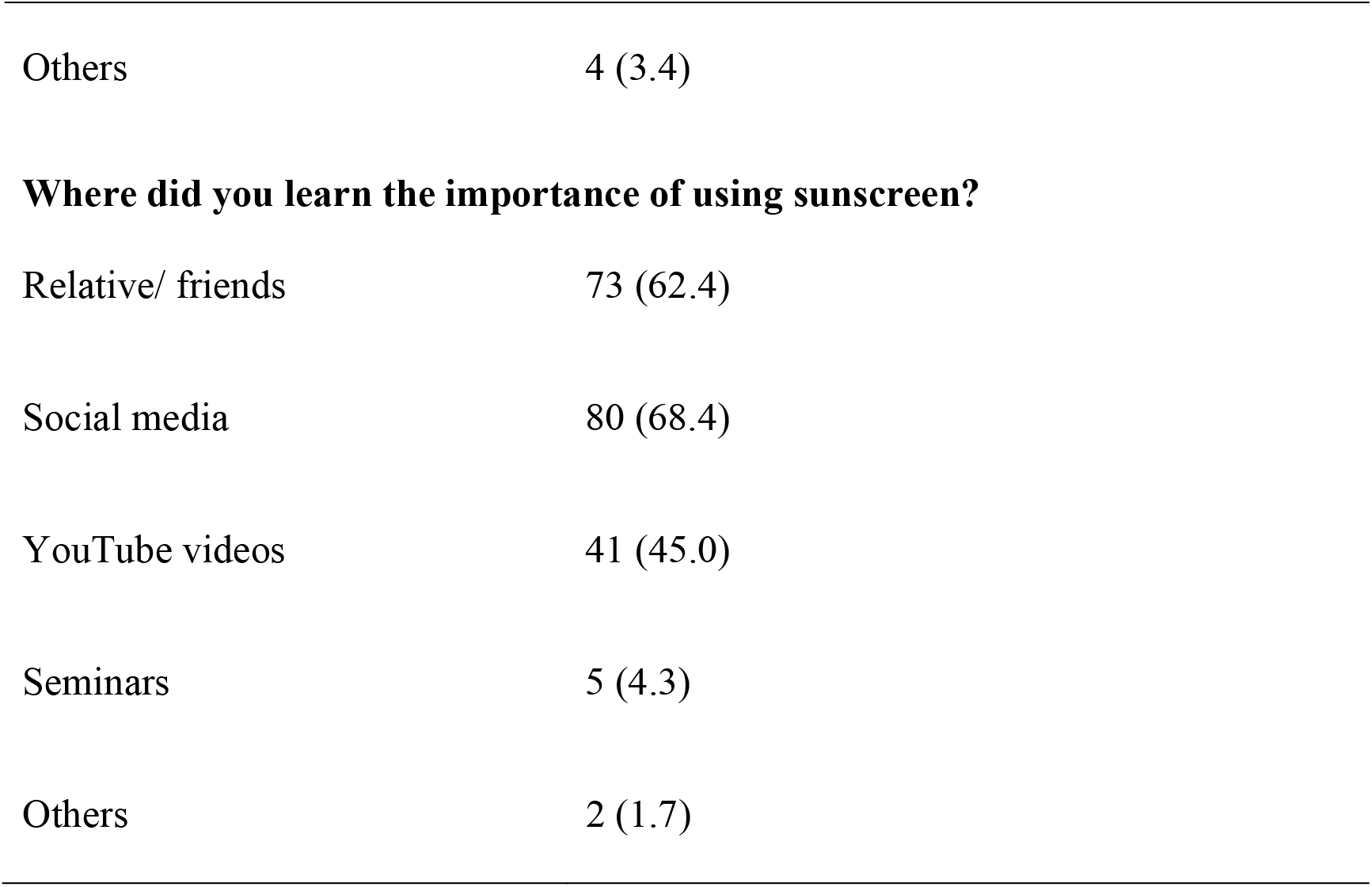
Practice of using sunscreen among medical students (n=117)

| Practice of using sunscreen | n (%) |
| --- | --- |
| **Do you use sunscreen every time you go out in the daylight?** |  |
| Always | 25 (21.3) |
| Mostly | 22 (18.8) |
| Sometimes | 17 (14.5) |
| Rarely | 29 (24.8%) |
| Never | 24 (20.5%) |
| **Is your sunscreen application a seasonal habit?** |  |
| Only sunny days | 29 (24.8) |
| All year round | 40 (34.2) |
| Rarely ever | 48 (41.0) |
| **What SPF of sunscreen do you use?** |  |
| 0-20 | 8 (6.8) |
| 20-50 | 29 (24.8) |
| 50+ | 49 (41.9) |
| BB or CC cream | 2 (1.7) |
| Does not matter | 29 (24.8%) |
| **How often do you reapply your sunscreen in a day?** |  |
| No reapplication | 105 (89.7) |
| Every hour | 2 (1.7) |
| Every few hours | 10 (8.5) |
| **When do you apply your sunscreen?** |  |
| Immediately before sun exposure | 30 (25.6) |
| 30 mins before sun exposure | 56 (47.9) |
| 1 hour before sun exposure | 12 (10.3) |
| 2 hours before sun exposure | 4 (3.4) |
| **What encourages you to wear sunscreen?** |  |
| Skin sensitive condition | 36 (30.8) |
| Doctor prescription | 5 (4.3) |
| Avoid tan/sunburn | 94 (80.3) |
| Protection against skin cancer | 59 (50.4) |
| Protection against aging | 39 (33.3) |
| Others | 3 (2.6) |
| Does not use sunscreen | 5 (4.3) |
| **What prompts you to avoid using sunscreen?** |  |
| Forgetting to apply | 81 (69.2) |
| Applying is a hassle | 39 (33.3) |
| Skin looks greasy / oily | 42 (35.9) |
| Costly | 16 (13.7) |
| It is not effective | 7 (6.0) |
| Others | 4 (3.4) |
| **Where did you learn the importance of using sunscreen?** |  |
| Relative/ friends | 73 (62.4) |
| Social media | 80 (68.4) |
| YouTube videos | 41 (45.0) |
| Seminars | 5 (4.3) |
| Others | 2 (1.7) |

## Notes

### Competing Interest Statement

The authors have declared no competing interest.

### Funding Statement

This study did not receive any funding.

### Author Declarations

The ethics approval to conduct this study was granted by the Research Ethics Committee, Faculty of Medicine, Manipal University College Malaysia (MUCM), Malaysia.

